# Deep neural networks allow expert-level brain meningioma detection, segmentation and improvement of current clinical practice

**DOI:** 10.1101/2021.05.11.21256429

**Authors:** Alessandro Boaro, Jakub R. Kaczmarzyk, Vasileios K. Kavouridis, Maya Harary, Marco Mammi, Hassan Dawood, Alice Shea, Elise Y. Cho, Parikshit Juvekar, Thomas Noh, Aakanksha Rana, Satrajit Ghosh, Omar Arnaout

## Abstract

Accurate brain meningioma detection, segmentation and volumetric assessment are critical for serial patient follow-up, surgical planning and monitoring response to treatment. Current gold standard of manual labeling is a time-consuming process, subject to inter-user variability. Fully-automated algorithms for meningioma detection and segmentation have the potential to bring volumetric analysis into clinical and research workflows by increasing accuracy and efficiency, reducing inter-user variability and saving time. Previous research has focused solely on segmentation tasks without assessment of impact and usability of deep learning solutions in clinical practice. Herein, we demonstrate a three-dimensional convolutional neural network (3D-CNN) that performs expert-level, automated meningioma segmentation and volume estimation on MRI scans. A 3D-CNN was initially trained by segmenting entire brain volumes using a dataset of 10,099 healthy brain MRIs. Using transfer learning, the network was then specifically trained on meningioma segmentation using 806 expert-labeled MRIs. The final model achieved a median performance of 88.2% reaching the spectrum of current inter-expert variability (82.6% - 91.6%) and compared to current workflows, reduced processing time by 99%. We demonstrate in a simulated clinical setting that a deep learning approach to meningioma segmentation is feasible, highly accurate and has the potential to improve current clinical practice.

## Introduction

Meningiomas are the most common primary intracranial neoplasms accounting for 37% of all primary brain tumors.^1^ They arise from the meninges and are benign in the vast majority of cases.^2^ Magnetic resonance imaging (MRI), and T1-weighted gadolinium contrast-enhanced sequences in particular, are the cornerstone of the diagnostic evaluation of meningiomas. A large number of scans are produced and examined throughout the life of any given patient for the purposes of initial diagnosis, clinical surveillance, surgical planning and post-operative assessment of residual tumor.^3-5^ Currently, assessment of meningioma imaging relies on manual techniques for tumor size and growth estimation, most commonly using unidimensional measurements in two or three orthogonal planes,^6^ which are often disregarded in favor of visual estimation of the tumor’s dimensions. Such approaches commonly lead to misjudgment of tumor growth and true dimensions and in some cases, due to their small dimensions, tumors are even completely missed. Manual volumetric segmentation is possible, typically with the help of third-party software, but is subject to considerable inter-rater variability^7^ and represents a time-consuming task that is often incompatible with the busy workflows of clinicians.

Advances in computing power and a gradual refinement of algorithm architectures have resulted in increased use of machine learning (ML) and deep learning (DL) techniques in healthcare.^8^ A specific class of DL architectures known as deep convolutional neural networks (CNN), in particular, has revolutionized imaging analysis.^9^ The impressive success of these networks in disparate tasks, such as diabetic retinopathy or skin cancer classification,^10,11^ has sparked an intense interest in employing them for other medical applications.^12^ Although there has been a considerable body of research focusing on the implementation of CNN segmentation algorithms for a number of brain pathologies, most notably glioblastoma,^13,14^ there is still a dearth of evidence in current literature on the application of end-to-end DL solutions for meningioma segmentation and management. Moreover, most brain tumor segmentation algorithms have focused on the algorithm’s labeling performance alone, and have not assessed the applicability and impact of such systems in real clinical scenarios.^15,16^ It is important to note that the readiness of a tumor segmentation algorithm for clinical use is not defined by 100% accuracy (which, though ideal, is extremely difficult to achieve), but rather by demonstrating performance at the same or higher level as the human experts who currently perform the task, taking into account the naturally occurring inter-expert variability. We therefore designed a study with the goal of developing a 3D-CNN algorithm able to automatically segment meningiomas from MRI scans at clinical expert level, and specifically offer objective measures of impact in a real-world clinical setting, focusing on the accuracy of tumor segmentation, volume estimation and time saving compared to current gold standards.

## Results

### Dataset creation and algorithm design

Our dataset consisted of 10,099 T1-weighted healthy brain MRIs, assembled from public and private sources, and 806 contrast-enhanced T1-weighted meningioma MRIs, representing 936 unique tumors, from the radiological repositories of two major academic hospitals under institutional review board approval. Details on data extraction, preprocessing and tumor labeling are found in the Methods section. In brief, tumor-containing MRIs were screened based on radiological or histological evidence of meningioma; high resolution brain scans were independently segmented by two experts (AB, VK) and reviewed by a third (AS). Sixty MRIs containing a total of 67 tumors were randomly held out as a test set; the rest of the database was used for training. A three-dimensional U-Net was used as the underlying architecture for our segmentation algorithm^17^ (Supplementary Fig. 1). The 10,099 normal brain MRIs served to train and validate the 3D-CNN for the task of brain extraction. The model was then fine-tuned via transfer learning to label brain meningiomas using 746 meningioma scans. The algorithm tumor-labeling performance was assessed with standard metrics of tumor segmentation performance (i.e., Dice score, Hausdorff distance). Three experts (VK, TN, PJ) independently segmented the tumors of the test set to provide a measure of inter-expert variability, while two experts (AB, AS) provided the time needed to perform manual segmentations.

### Algorithm performance

The tumors in the test set had a mean volume of 13.11 cc (range 0.37 - 85.0 cc) and all intracranial locations commonly harboring meningiomas were adequately represented: cranial vault (56.7%), skull base (25.3%), falx (13.4%), posterior fossa (4.5%)(Supplementary Fig. 2 and Supplementary Fig. 3). Five MRIs contained two tumors and one MRI contained three. The final model achieved a mean tumor segmentation Dice score of 85.2 % (mean Hausdorff = 8.8 mm; mean average Hausdorff distance = 0.4) and a median of 88.2% (median Hausdorff = 5.0 mm; median average Hausdorff distance = 0.2 mm) on the entire test set, comparable to the inter-expert variability in segmenting the same tumors with means ranging from 80.0 % to 90.3 % (Hausdorff distance means range = 5.5 to 14.6 mm; Average Hausdorff distance means range = 0.1 to 2.1 mm) and medians between 82.6% to 91.5 % (Hausdorff distance median range = 4.1 to 9.2 mm; Average Hausdorff distance median range = 0.1 to 0.2) (Table 1 and Supplementary Tables 1 and 2). With regards to detection ability, the model missed two small tumors (volume < 1cc) whereas two out of three independent experts missed four each (sensitivities: model: 97%, VK:100%, TN:94%, PJ%94%). On the other hand, the algorithm segmented additional 18 small vascular structures due to the similarity in contrast uptake and rounded shape whereas no expert segmented any normal anatomical structures.

We identified the patients who would be most likely to benefit from the use of a segmentation algorithm, i.e. those with the need of close radiological monitoring or higher chance of undergoing surgical resection, as those with tumors larger than 2cc; this cutoff is significantly stricter compared to the size of a clinically relevant meningioma based on the literature (∼4 cc).^18-20^ In this group of 41 patients, model and experts obtained equivalent performance, with the model demonstrating a mean tumor segmentation Dice score of 87.4 % (median 89.5) (mean and median Hausdorff distance = 9.9, 5.9 mm; mean and median average Hausdorff distance = 0.3, 0.1)and an inter-expert variability ranging between 84.1 and 91.2% (median range: 86.9 - 92.5) (Table 1, Fig. 1). Dice provides a measure of volumetric similarity^.41^ Hausdorff distance measures boundary similarity and is the greatest distance from the boundary of one object to the boundary of another object^.42^ Average Hausdorff distance is similar to Hausdorff distance but is less susceptible to outliers.^43^ There was a significantly positive correlation between tumor Dice scores and tumor volume with a steep performance increase in the 0-2 cc volume range (r=0.61, p<0.001, Fig. 2a).

**Table 1.** Algorithm performance and inter-expert variability. Comparison of tumor segmentation performance (Dice scores) between algorithm output, ground truth and clinical experts (Expert_1(VK), Expert_2(TN), Expert_3(PJ)), expressed as mean and median for all tumors and tumors of volume >2cc.

|  | **> 2cc tumors** |  | **All tumors** |  |
| --- | --- | --- | --- | --- |
| Pairs for comparison | Mean Dice score (%) | Median Dice Score (%) | Mean Dice score (%) | Median Dice Score (%) |
| **Performance** |  |  |  |  |
| Model/Ground | 87.7 | 89.6 | 84 | 88.2 |
| Model/Expert_1 | 86.9 | 88.7 | 82.7 | 86.4 |
| Model/Expert_2 | 87 | 90.2 | 83.6 | 88 |
| Model/Expert_3 | 85.2 | 89.3 | 80 | 85.1 |
| **Inter-Expert Variability** |  |  |  |  |
| Ground/Expert_1 | 87.6 | 89.1 | 85.7 | 86.8 |
| Ground/Expert_2 | 91.2 | 92.5 | 89.9 | 91.6 |
| Ground/Expert_3 | 89.5 | 90.5 | 86 | 89.5 |
| Expert_1/Expert_2 | 86.9 | 88.9 | 84.6 | 86 |
| Expert_1/Expert_3 | 84.1 | 86.9 | 79.5 | 82.6 |
| Expert_2/Expert_3 | 90.1 | 92.7 | 87 | 89.7 |

*Ground:* Ground truth

**Fig. 1.**
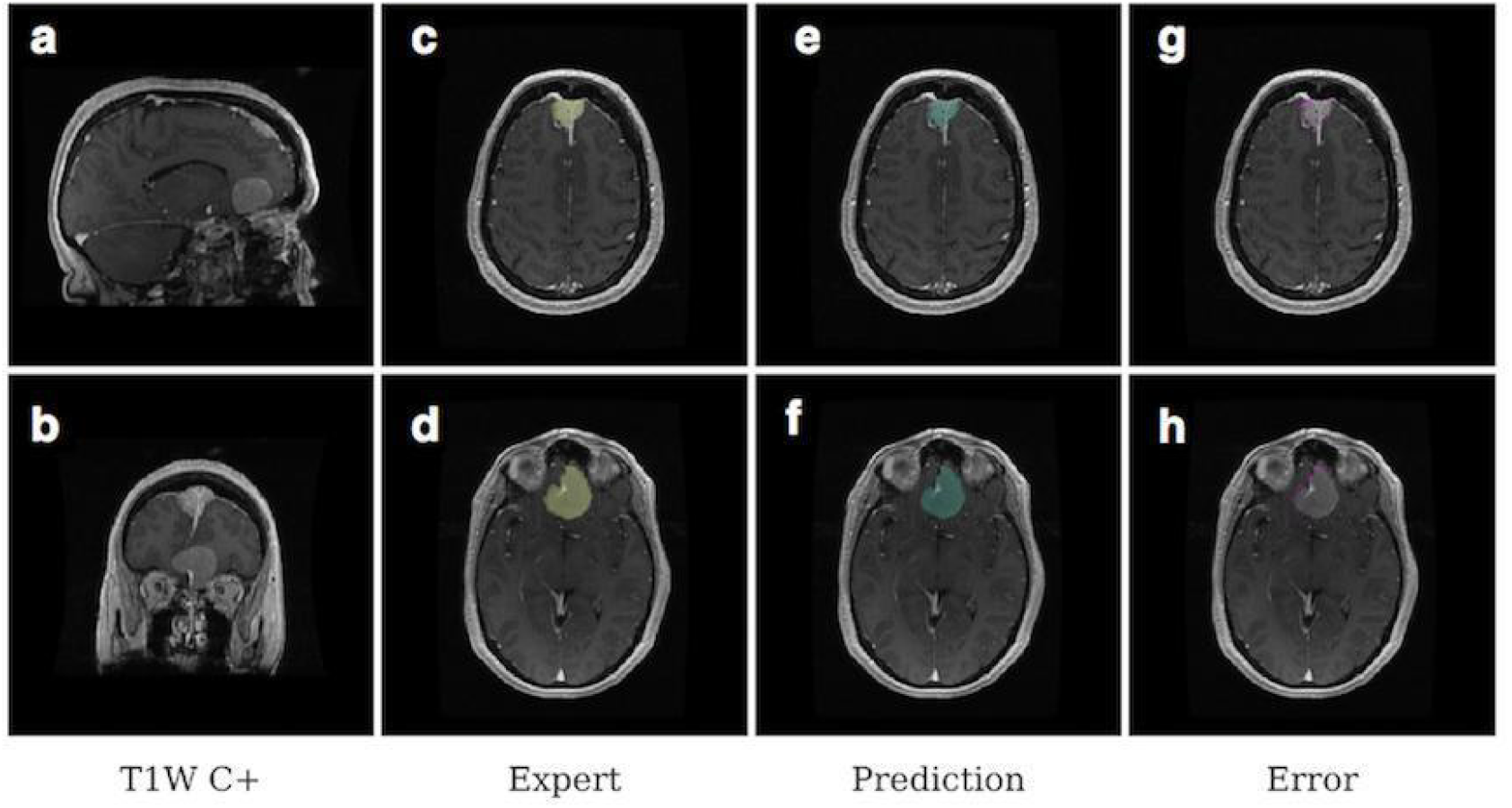
Example of meningioma segmentation algorithm output. Sagittal and coronal views (**a** and **b**) of a brain MRI scan containing two distinct meningiomas, one located in the convexity at the midline, the other located on the anterior skull base. Display of expert label vs computer-generated segmentation respectively of the meningioma of the convexity (**c** and **e**) and the meningioma of the skull base (**d** and **f**). Display of the mismatch between the expert label and the computer-generated segmentation on the meningioma of the convexity (**g**) and the meningioma of the skull base (**h**).

**Fig. 2.**
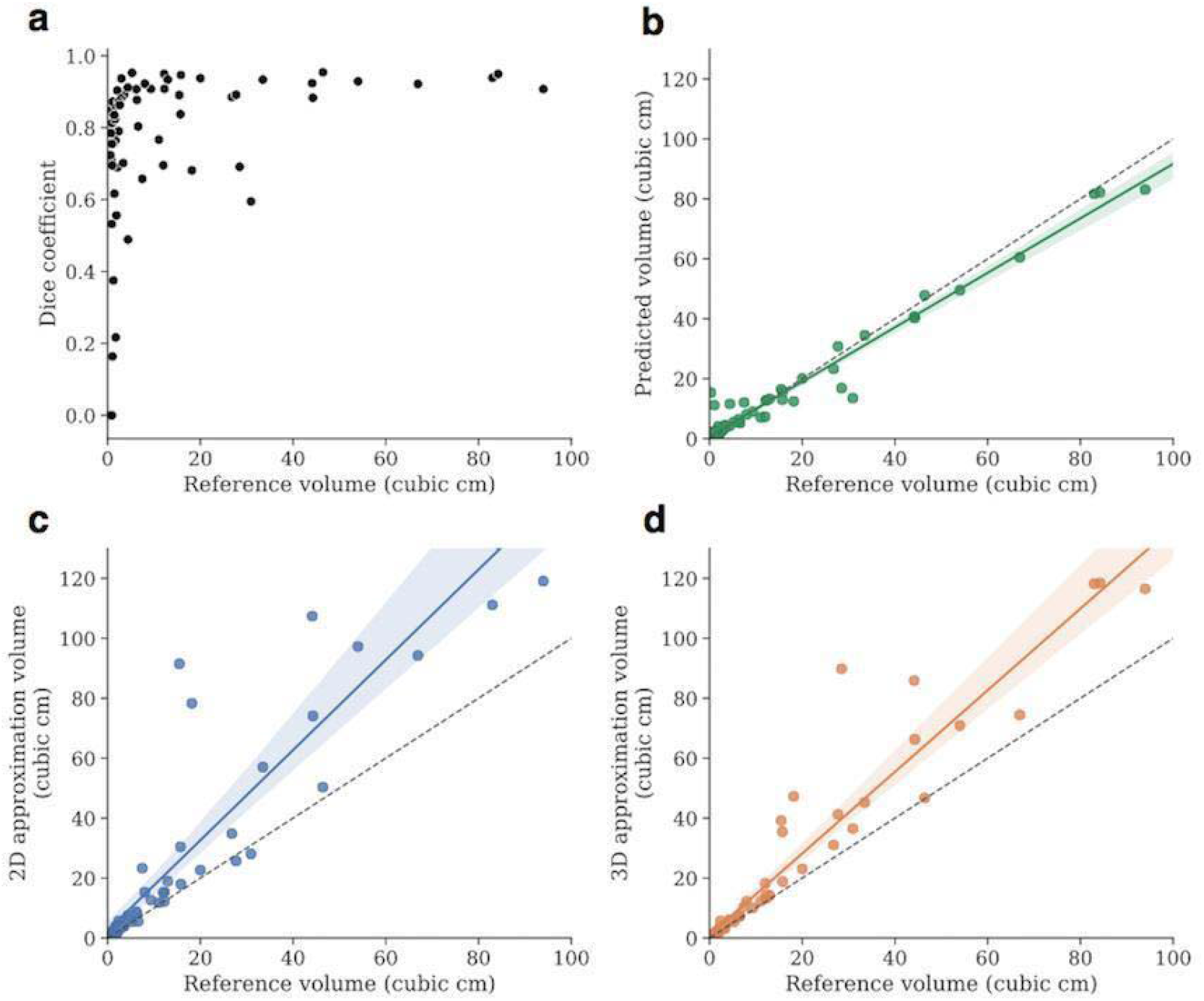
Algorithm’s tumor segmentation and volume estimation accuracy. (**a**) Scatter plot showing the algorithm’s segmentation performance expressed as Dice scores on the test set as a function of the tumor volume. The Dice score correlated with the size of the tumor, quickly reaching a mean of 0.87 and median of 0.89 for tumors > 2cc. (**b, c, d**) Volume estimations by the algorithm, and the 2-D and 3-D traditional estimation techniques, respectively. The algorithm’s predicted volumes constitute almost perfect approximations of the real tumor volumes with a correlation of 0.98, p<0.001 (**b**), whereas the 2D and 3D techniques present lower values of respectively 0.88 (**c**) and 0.96 (d) p<0.001, with evidence of overestimation.

### Measures of clinical impact

We designed a simulated clinical scenario to compare volume estimation accuracy and segmentation time based on current practice versus the use of our automated algorithm. In brief, two certified experts, one neuro-radiologist (AS) and one neurosurgeon (AB), were asked to manually segment each tumor in the test set while timing themselves, and tumor volumes were calculated with 2D/3D estimation techniques currently used in clinical practice (setting and details reported in the Methods section). Notably, the incorporation of our algorithm reduced segmentation time by 99% (two seconds per segmentation on average, p<0.001, Table 2), and produced tumor volume calculations with an almost perfect correlation with the expert manual segmentations (r=0.98, p<0.001, Fig. 2b, Supplementary Tables 3 and 4), significantly more accurate compared to the gold standard 2D and 3D volume estimation techniques (r=0.88 p<0.001 and r=0.96, p<0.001, respectively, Fig. 2c, 2d, Supplementary Tables 3 and 4). Currently used 2D/3D estimation techniques were found to overestimate tumor volume by a minimum of 1.5 to a maximum of 7 times compared to the algorithm-based calculation (Supplementary Tables 4 and 5).

**Table 2.** Manual versus automated tumor segmentation times. Comparison between the average time needed by experts **(Manual 1** and **Manual 2)** and our algorithm (**Automated**) to produce high-quality tumor segmentations. The algorithm saved 98% of time on average and the difference between the algorithm and each expert in each tumor group reached significance (p<0.001).

|  | **Mean (s)** | **SD (s)** | **Time reduction (%)** | **p value** |
| --- | --- | --- | --- | --- |
| **All tumors** |  |  |  |  |
| Manual 1 | 142.4 | 93.9 | 98.7 | **< 0.001** |
| Manual 2 | 299.8 | 274.9 | 99.4 | **< 0.001** |
| Automated | 1.88 | 0.001 | - | ref |
| **>2cc** |  |  |  |  |
| Manual 1 | 178.2 | 96.2 | 98.9 | **< 0.001** |
| Manual 2 | 391.5 | 296.9 | 99.5 | **< 0.001** |
| Automated | 1.88 | 0.001 | - | ref |
| **<2cc** |  |  |  |  |
| Manual 1 | 70.9 | 17.3 | 97.3 | **< 0.001** |
| Manual 2 | 116.4 | 23.6 | 98.3 | **< 0.001** |
| Automated | 1.88 | 0.001 | - | ref |

s: seconds, ref: reference

## Discussion

Here, we demonstrate the development of an end-to-end DL solution for automated brain meningioma segmentation. The system was trained on the most heterogeneous MRI dataset of expert-labeled meningiomas available in the literature and found to have excellent detection and segmentation performance compared to human experts. The 3D-CNN algorithm performed at the same level of human experts while outperforming comparable automated or semiautomated segmentation algorithms.^21-23^ Specifically, the model has well surpassed current gold standards in tumor volume estimation while bringing the time needed for tumor segmentation virtually to zero. The Dice scores suggest that there is a high degree of overlap between the reference and the model output, and the relatively low Hausdorff scores indicate that the surfaces of the model outputs are similar to the surfaces of the reference labels. With regards to detection, while model’s ability to correctly detect tumors was comparable to human experts, the challenge remains to distinguish between tumoral and vascular structures due to the similarity in contrast uptake and rounded shape.

Although others have described segmentation algorithms with good performance, these have been based on smaller datasets, which either only include patients with larger tumors (average volume 30 cc) and/or those undergoing surgical resection,^22,23^ thereby excluding almost 50% of patients affected by meningiomas who normally undergo conservative MRI surveillance.^24^ Such inclusion criteria, alongside complex pre-processing pipelines, limit the implementation of these algorithms in a real-world clinical population. In order to optimize the applicability and generalizability of our system, we (1) limited the artificial curation of the dataset by keeping image pre-processing to a minimum, simply ensuring standardization between MRI scans produced by different machines (more details in the Methods section), included MRIs with no restrictions on tumor number, size, and location and (3) included patients who underwent both conservative management or surgical treatment. These features make our dataset more representative of the variability inherent in the meningioma patient population. In addition, we report a 3D U-Net pre-trained on 10,000 non-clinical T1-weighted MRI scans. The scale of the pre-training data makes our 3D U-Net a viable candidate for fine-tuning, and to the knowledge of the authors, this is the largest-scale 3D U-Net that is publicly available for brain extraction and fine-tuning. This greatly reduces the barriers to deep learning for brain imaging groups, which often operate in low-data regimes.

Some important limitations of this study should be highlighted: the inclusion of single, pre-operative scans alone didn’t allow the evaluation of post-operative residuals, tumor recurrence or tumor growth; with regards to tumor detection ability, while absolute numbers and sensitivity were reported, it was not possible to determine specificity at the tumoral level due to the difficulty to define the entirety of true negatives (normal anatomical structures); the real advantage in terms of work schedule streamlining would necessitate the inclusion of the algorithm in the software pipeline of the hospital informatic system which was not possible at the stage of the current study; while designed as a clinical simulation, the present study is retrospective in nature and will necessitate of prospective validation before considering real-world clinical applicability.

Despite a growing interest in the application of DL segmentation algorithms to medical imaging, few studies have evaluated the feasibility of applying these tools in daily clinical scenarios.^25,26^ Our goal was to bridge the gap between quantitative research and clinical practice. Thus, we chose to evaluate our system not purely based on its performance in the segmentation task but also with real-world metrics. With a tumor detection and segmentation performance comparable to that obtained by clinical experts, a superior volumetric calculation accuracy compared to current 2D/3D estimations and a rapid, completely automated process, our algorithm has the potential to improve brain meningioma workflow management (Supplementary Fig. 4).. It could allow for more accurate tumor information acquisition and growth monitoring which would in turn improve clinical decision-making as well as monitoring of treatment response.

As with all DL algorithms, performance and generalizability improve with enrichment of the dataset. In the context of this work, this will be accomplished through validation on data from other institutions to help control for any institutional biases in terms of MR acquisition and patient population. An additional step towards full implementation of our tool into clinical work-flow would entail the evaluation of the algorithm performance in segmenting residual or recurrent meningioma in the setting of early and late post-operative changes. Both these steps to optimize the algorithm are currently underway.

In addition to the relevance of a fully automated DL segmentation tool, such models are important upstream components in more complicated ML pipelines that have the potential to assist in predicting parameters such as tumor consistency, histological grade, growth trajectory and probability of recurrence even before a therapeutic plan is made and discussed with the patient.^27-29^ Such systems could enhance clinical decision-making strategies, patient counseling and follow-up in unprecedented ways. To ensure that the development of DL algorithms for medical imaging analysis is more translational in nature, the impact of these systems needs to be validated on simulations of actual clinical tasks in the setting of a well-designed clinical trial. This will allow the selection and eventual implementation of the best performing algorithms which, with the necessary consideration of patient privacy and safety and the clearance provided by the dedicated certification and standardization pathways,^30,31^ will be able to effectively advance medical care and improve patient outcomes.^32,33^ Most important, the medical community will have to be an active participant in this process, working alongside computer scientists in order to steer the development towards the most relevant clinical questions, guarantee the rigorous validation of these algorithms and their seamless integration in clinical practice.

## Methods

### Image acquisition and preprocessing

#### Healthy brain dataset

A dataset of 10,099 high-resolution, T1-weighted MRI scans of normal human brains was assembled from public and private sources.^34,35^ This dataset contained images from a heterogenous human population and of varying acquisition quality. All scans were conformed to have 256 slices in each dimension and 1mm^3^ isotropic voxels using the image processing software FreeSurfer.^36^ FreeSurfer’s recon-all tool was used to generate parcellations and segmentations for brain structures in each scan. The dataset was separated into a training/validation set.

#### Meningioma dataset

We screened all adult patients diagnosed with intracranial meningioma in two major US academic hospitals during the time period 2004-2018 for inclusion in our study. Patients with no evidence of meningioma on MRI, unavailable pre-surgical MRI scans, radiation-induced meningioma, unavailable contrast-enhanced images, or no high-resolution sequences were excluded, resulting in a final cohort of pre-surgery 806 exams, containing a total of 936 tumors. The exams were downloaded in DICOM format and stored in an institutionally protected, HIPAA compliant shared drive. Per protocol, high resolution (at least 100 slices on axial plane), T1 contrast-enhanced sequences were identified from our final cohort and converted to NIfTI format in order to ensure patient de-identification. The meningioma dataset was divided into a training/validation set (746 scans) and a test dataset of 60 exams containing a total of 67 tumors. The test set was constructed in order to ensure adequate representation of all the most relevant tumor locations, sizes and shapes. All relevant ethical regulations were followed as part of the conduct of this study. We operated under institutional review board approval by the Partners Human Research Committee who granted permission for this research project (protocol number 2015P002352).

### Ground truth tumor labeling

We used the open-source image-processing software 3D Slicer to produce expert-level meningioma segmentations. 3D Slicer provides both manual and semi-automated labeling tools that the experts could choose from to perform the task. All tumors were segmented by two experts (AB is a fully trained neurosurgeon with 5 years’ experience in brain tumor segmentation; VK is a neurosurgery resident with extensive computational background and 2 years’ experience in brain tumor segmentation) separately and inter-rater variability was calculated. The labels were reviewed by a third expert (AS is a fully trained neuro-radiologist with 5 years of clinical experience in evaluation of location, extension and features of brain tumor lesions) in order to produce the final dataset. The single final ‘ground truth’ was determined after evaluating both segmentations sets. In case of tumors of particular complexity, all three experts independently segmented the lesion and jointly discussed the result in order to come up with a shared agreement on the ground truth. The segmentations were stored in an institutional shared drive and named using progressive numeration; the link between each exam and the corresponding patient was securely stored in a separate file. Location, size and number of tumors per scan were stored in a dedicated spreadsheet for subsequent analysis.

### CNN model design and evaluation

#### Brain extraction network

A standard three-dimensional U-Net was used as the underlying architecture for our segmentation algorithm.^20^ Olaf Ronneberger et al. originally designed the UNet architecture for a bio-medical image segmentation problem. It has been observed to be more successful than other conventional CNN models, in terms of architecture and in terms pixel/voxel-based segmentations from convolutional neural network layers.^37^

The U-Net architecture is primarily composed of an encoder and a decoder. The encoder can be seen as a contraction path - a stack of convolutional and max pooling layers which captures and encodes the contextual information present in the input. The decoder is an expansion path that is used to decode the accurate localization using the learned feature mapping from the input and up-convolutional layers (Supplementary Fig. 1).

The model was initially trained with random weight initialization for a brain-extraction task on the dataset of 10,000 T1-weighted MRIs of healthy brains. To improve generalizability and robustness of the network, each pair of MRI and labels had a fifty-percent chance of being augmented using random rotation and translation (the transformation for pairs of features and labels was the same). The augmented MRIs were interpolated tri-linearly, and the augmented reference labels were interpolated using nearest neighbor. The voxels of each MRI scan were standard scored. Due to the GPU computational limitations, the MRI scans and labels were separated into eight non-overlapping cubes of equal size per volume. Model training was done on these cubes in batches of six. The model was trained using the Adam optimizer^38^ with an initial learning rate of 0.00001 and Jaccard loss for five epochs across three NVIDIA GTX 1080Ti graphical processing units (GPUs). Each batch was distributed evenly to each GPU. The model was implemented using the Nobrainer framework, which wraps the Keras API of TensorFlow.^39^ The random augmentation and interpolations were implemented in the Nobrainer framework using TensorFlow. Dice similarity scores, Hausdorff distance and average distance were calculated and used to evaluate the algorithm’s performance in brain extraction. Dice was calculated using NumPy, and Hausdorff was calculated using SciPy. Average Hausdorff distances were calculated using the EvaluateSegmentation tool described by Taha and Hanbury.^40^ The model is publicly available in the Keras format at https://github.com/neuronets/nobrainer-models/releases/tag/0.1.

#### Meningioma extraction network

The refined brain-extraction network was re-trained for the specific task of meningioma segmentation. The meningioma training/validation dataset underwent the same preprocessing pipeline as the normal brain scans, with the only difference that the tumor-containing scans were not randomly augmented. The MRI scans and labels were separated into eight non-overlapping cubes of equal size per volume. The cubes of the MRI scans that did not contain any meningioma voxels were excluded from the training set. To prevent large deviations from the pre-trained weights, L2 regularization with coefficient 0.001 was applied to the weights of each layer and a low initial learning rate of 0.00001 was used. The model was trained using the Adam optimizer^38^ and the Jaccard loss for 194 epochs with a batch size of two cubes using a single NVIDIA GTX 1080Ti GPU. Validation on whole MRIs was performed by standard-scoring the MRI voxel values, separating the MRI into eight non-overlapping cubes of equal size, running a forward pass of the model on each cube, and combining the eight cubes of predictions into a complete volume. The algorithm’s segmentation performance was assessed by computing the Dice coefficient between the tumor model prediction and the expert label for each MRI in the test set.

An experimental attempt was also done to train a model using the meningioma dataset alone from random weight initialization with no implementation of transfer learning. The attempt was abandoned at its early stage due to its clear inferiority in the segmentation performance compared to the algorithm adopting transfer learning described in this work.

### Expert segmentation variability range

Three different experts (VK, TN is a fully trained neurosurgeon and fellow in advanced brain imaging, PJ is a research fellow in advanced brain imaging with computational background and 2 years of experience in brain tumor segmentation and brain tractography) were given the task to independently segment the tumors of the test set in order to obtain a segmentation variability range. Mean and median tumor segmentation results were calculated for each expert and used to create a range of expert segmentation variability compatible with a real-world clinical scenario. The model performance was compared to the obtained range of values.

### Clinical simulation

To obtain the most clinically meaningful metrics as to whether our deep learning segmentation algorithm can effectively impact the current clinical practice by improving physicians’ accuracy in calculating meningioma volume and reducing the time needed to produce high-quality, readily usable tumor segmentations, we simulated a clinical scenario. In the trial we put the algorithm against currently accepted methods of tumor volume assessment and measured the difference in time between clinical experts and our algorithm to produce volumetric tumor segmentations (described below).

#### Tumor volume estimation

We compared our algorithm’s volume estimation accuracy to the two most widely employed tumor volume estimation techniques, based on 2-dimensional (2D) and 3-dimensional (3D) tumor volume calculation. The 2D technique is based on the selection of the MRI axial slice containing the largest tumor cross-section, followed by measurement of the tumor’s two largest diameters, called tumor length and tumor width, respectively. The volume is then calculated as follows:

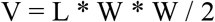

where V is the estimated tumor volume, L is the tumor length and W is the tumor width. In the 3D-based technique, three slices containing the largest tumor cross-section in the three traditional MRI visualization planes (sagittal, axial, coronal) are chosen and the largest diameter on each plane is measured, called length, width and height of the tumor, respectively. The volume is then calculated as follows:

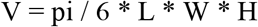

Where V is the tumor volume, L is the largest sagittal diameter, W is the largest axial diameter, and H is the largest coronal diameter. These two techniques were applied to each MRI scan of the test set through a dedicated software tool for the detection of the longest diameter in each of the orthogonal planes. The tumor volumes from the algorithm’s output labels on the same test set were calculated as well. The volumes resulting from these three methods were compared to those obtained from the ground truth labels and the correlation between each pair of volumes was calculated using the r statistic (2D to ground truth, 3D to ground truth, algorithm-volume to ground truth).

#### Segmentation time analysis

Two certified experts (AB and AS) who regularly work with brain MRIs containing meningiomas for diagnostic, surgical planning and patient follow-up purposes were given the task of timing themselves in producing an accurate, readily usable meningioma label starting from an unedited MRI scan for each exam included in the test set. They were instructed to open the exam and work on the segmentation of the lesion as they would normally do, with the software tools they were most comfortable with (brush, region-growing tool, pencil and eraser). Once they finished with an exam, they would save the file, close it, and proceed to the next one. Each event was timed from the moment the exam was opened and ready in the viewing software (i.e., 3D Slicer) until the segmentation was considered complete by the expert. Concurrently, our algorithm was used to produce the tumor labels of the same MRI scans and the related run time was stored. The average time difference was calculated between each expert and the algorithm. Given the non-normal distribution of the three sets of measurements, the Wilcoxon signed-rank test was applied to assess statistical significance.

## Data Availability

Data Availability
The majority of the healthy brain MRI dataset used and analyzed during the current study is publicly available at https://sensein.github.io/open-data-processing/. Restrictions apply to part of the healthy brain MRI dataset and the whole meningioma MRI dataset as derived from private, institutional databases and contain information that could enable patient identification. The authors will consider request to access to training and testing data on individual basis. Any data use will be restricted to non-commercial research purposes, and the data will only be made available upon execution of appropriate data use agreements.
Code availability
As reported in the Methods section of the manuscript, the code for the algorithm development and evaluation is publicly available at: https://github.com/neuronets/ams and the pretrained model is available at https://github.com/neuronets/trainedmodels/tree/master/neuronets/ams/0.1.0.
 

https://sensein.github.io/open-data-processing/.

https://github.com/neuronets/ams

https://github.com/neuronets/trainedmodels/tree/master/neuronets/ams/0.1.0.

## Acknowledgements

J.R.K was partially supported by R01EB020740, P41EB019936, and T32GM008444. S.G. was partially supported by R01EB020740, P41EB019936, and RF1MH121885. A.R. was supported by RF1MH121885. Shreya Chawla, Sharmila Devi and Ali Ansaripour from the Computational Neuroscience Outcomes Center provided support for data extraction and pre-processing. We are grateful to Margherita De Marzio and Enrico Maiorino for their independent and precious feedback.

## Author Contributions

A.B., S.G., O.A. contributed to the study design. A.B., V.K.K., M.H., H.D. were responsible for patient selection and data collection. M.M., Z.Y.C. contributed to data extraction and pre-processing. J.R.K. and S.G. were responsible for experiments and data analysis and created the figures. A.B., V.K.K., A.S., P.J., T.N. contributed to the analysis. A.B., J.R.K., V.K.K., M.H., A.R. contributed to the data interpretation and to the writing. O.A. and S.G. were the senior supervisors of the project.

## Data Availability

The majority of the healthy brain MRI dataset used and analyzed during the current study is publicly available at https://sensein.github.io/open-data-processing/. Restrictions apply to part of the healthy brain MRI dataset and the whole meningioma MRI dataset as derived from private, institutional databases and contain information that could enable patient identification. The authors will consider request to access to training and testing data on individual basis. Any data use will be restricted to non-commercial research purposes, and the data will only be made available upon execution of appropriate data use agreements.

## Code availability

As reported in the Methods section of the manuscript, the code for the algorithm development and evaluation is publicly available at: https://github.com/neuronets/nobrainer-models/releases/tag/0.1.

**Supplementary Fig. 1. Deep Neural Network Architecture**. The architecture of our deep neural network consisted of a 3D-UNet structured with an encoder and a decoder arm. The network accepts 3D MRI data as input and outputs a 3D segmentation map.

**Supplementary Fig. 2. Tumor location distribution**. Bar plot showing tumor location distribution (% of the total) in the whole meningioma dataset and test set. All the main anatomical locations are adequately represented both in the general dataset and in the test set.

**Supplementary Fig. 3.** *Left*. Histogram showing the volume distribution of meningiomas in the training set. *Right*. Histogram showing the volume distribution of meningiomas in the test set.

**Supplementary Fig. 4. Current clinical workflow and clinical workflow implementing automatic tumor segmentation**. *Red light*: Volumetric tumor mask is not available; *Green light*: Volumetric tumor mask is available. Above: Volumetric tumor segmentation is currently implemented manually and only for surgical planning purpose, while tumor shape and volume information are not routinely available. Below: The implementation of an automated, expert-level tumor segmentation algorithm provides a tumor mask with accurate shape and volume information at each point of patient care, from the day of the first MRI scan in the radiology suite to the operating room, to each radiologic and clinical follow-up patient encounter.

