## Supplementary Table 1 for "Deep neural networks allow expert-level brain meningioma detection, segmentation and improvement of current clinical practice"

**Supplementary Table 1. Accuracy measures (Dice scores) between algorithm predictions and experts’ segmentations**. The table includes accuracy measures (Dice scores) for each MRI in the test set, used to calculate mean and median accuracy measures between algorithm predictions (**Prediction**) and ground truth (**Ground**), and to evaluate inter-expert variability between clinical experts (**Expert_1, Expert_2, Expert_3**) and ground truth and within clinical experts.

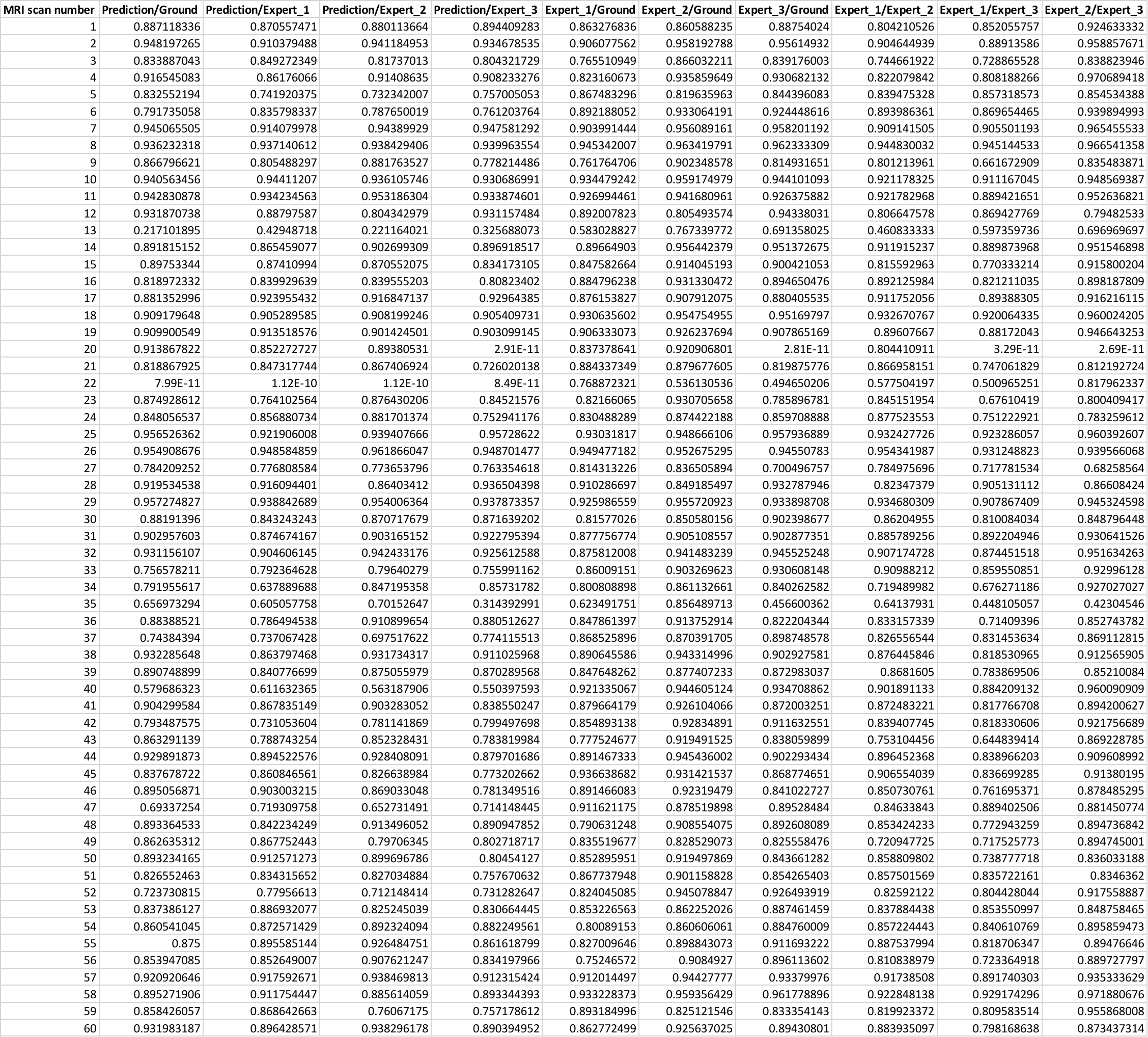
