## Supplementary Table 5 for "Deep neural networks allow expert-level brain meningioma detection, segmentation and improvement of current clinical practice"

**Supplementary Table 5. Summary statistics of over- and under-estimation of tumor volume.** The summary statistics compare the volume of the ground truth labels to other labeling techniques. The “overestimation” column contains statistics for volumes in which the labeling technique over-estimated the volume relative to the ground truth, and *vice versa* for the “underestimation” column. N indicates the number of MRIs used in the summary statistics. The summary statistics are in units of cubic centimeters.

| **Ground truth** | **Overestimation** | | **Underestimation** | |
| --- | --- | --- | --- | --- |
|  | **N** | **Mean (Min - Max) (cc)** | **N** | **Mean (Min - Max) (cc)** |
| Model output | 14 | 0.37 (0.01 - 1.22) | 46 | 2.37 (0.02-18.03) |
| Expert_1 | 1 | 0.13 (0.13 - 0.13) | 59 | 2.30 (0.05 - 13.69) |
| Expert_2 | 23 | 0.74 (0.01 - 3.20) | 37 | 1.17 (0.00 - 9.16) |
| Expert_3 | 47 | 0.86 (0.03 - 4.62) | 13 | 2.08 (0.07 - 7.53) |
| 2D estimation | 52 | 12.23 (0.02 - 110.95) | 8 | 0.85 (0.03 - 2.85) |
| 3D estimation | 56 | 7.09 (0.01 - 61.37) | 4 | 0.13 (0.02 - 0.20) |
