## Supplementary Table 4 for "Deep neural networks allow expert-level brain meningioma detection, segmentation and improvement of current clinical practice"

**Supplementary Table 4. Average absolute tumor volume differences between techniques**. The table presents the average tumor volume differences between ground truth and respectively the model output, expert manual segmentations (**Expert_1, Expert_2, Expert_3**) and 2D/3D volume estimation techniques (**2D estimation, 3D estimation**).

| Ground truth | Average absolute volume difference (cc) | | |
| --- | --- | --- | --- |
| VS | All tumors | >=2cc tumors | <2cc tumors |
| Model output | 1.90 | 2.66 | 0.27 |
| Expert_1 | 2.27 | 3.17 | 0.32 |
| Expert_2 | 1.01 | 1.43 | 0.08 |
| Expert_3 | 1.12 | 1.52 | 0.26 |
| 2D estimation | 10.72 | 15.44 | 0.53 |
| 3D estimation | 6.62 | 9.54 | 0.33 |
