## Supplementary Table 3 for "Deep neural networks allow expert-level brain meningioma detection, segmentation and improvement of current clinical practice"

**Supplementary Table 3. Tumor volume values**. Tumor volume values (in cc) for each MRI in the test set, calculated for ground truth (**Ground**), algorithm prediction (**Predicted**), 2D and 3D estimation techniques (**2D estimation, 3D estimation**), clinical experts’ segmentations (**Expert_1, Expert_2, Expert_3**). These measures were used to evaluate differences in tumor volume calculation between algorithm, clinical experts and estimation techniques.

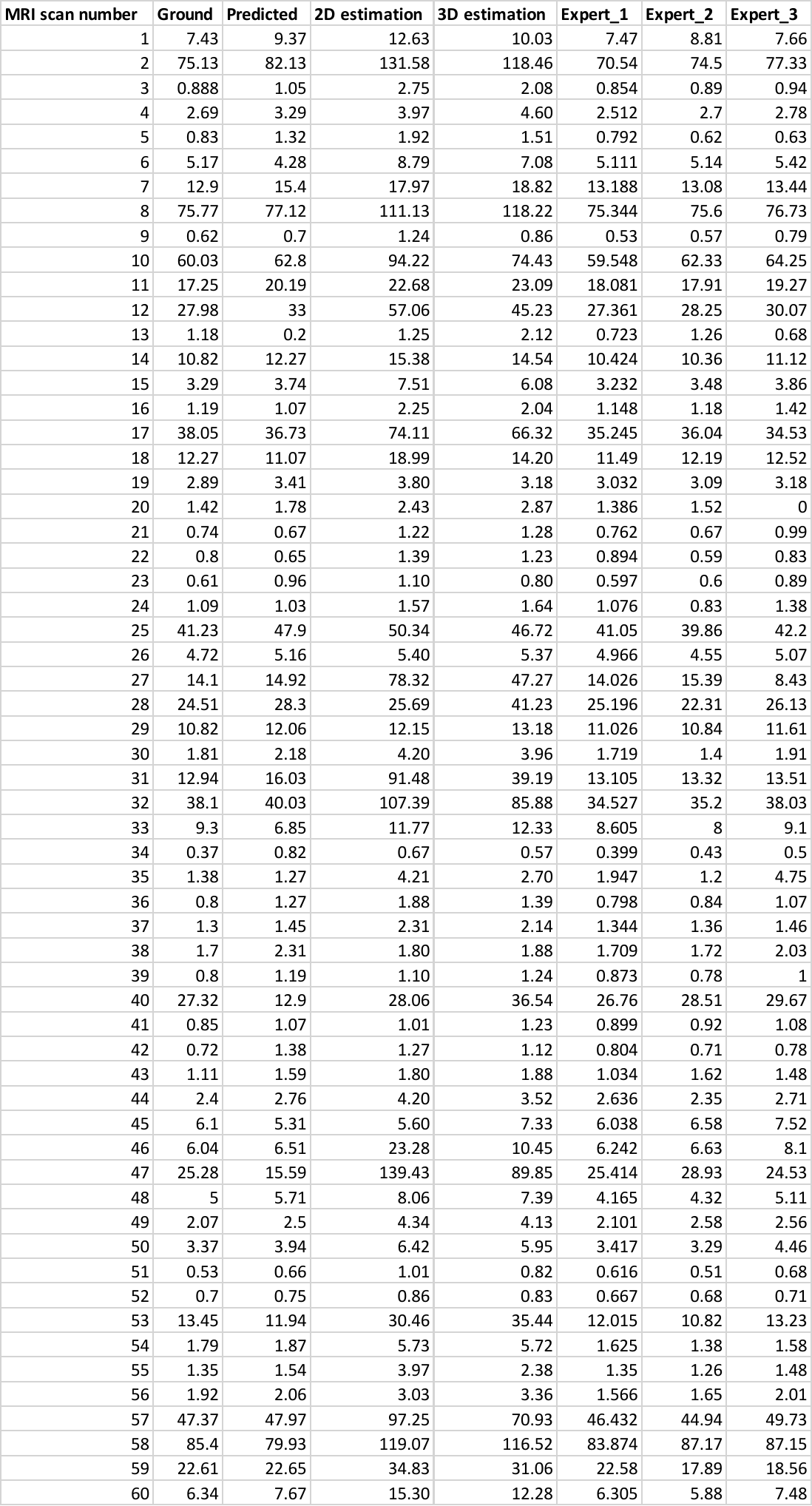
