## Supplementary figures and images for "Deep neural networks allow expert-level brain meningioma detection, segmentation and improvement of current clinical practice"

### Supplementary Figure 1

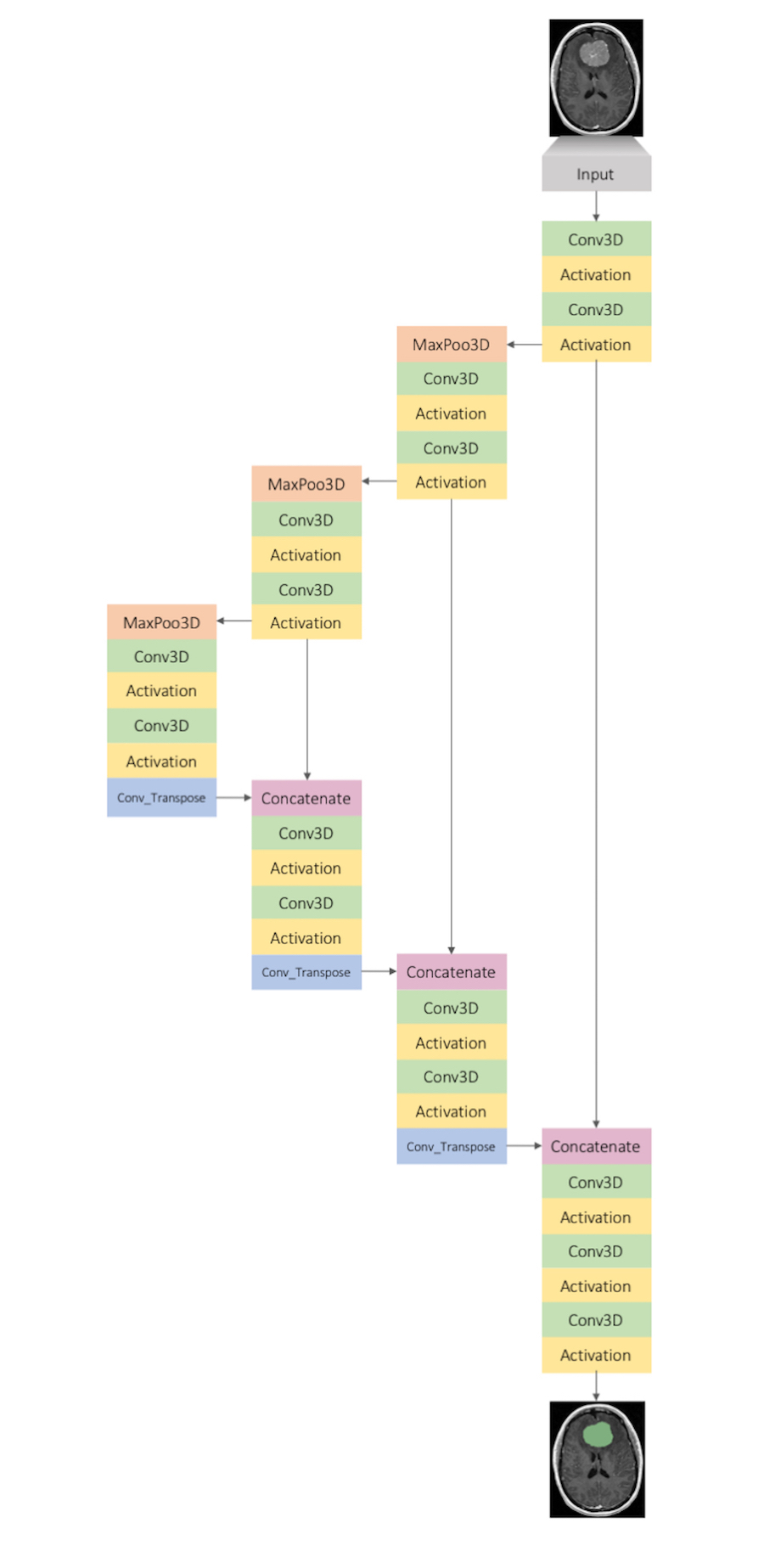

### Supplementary Figure 2

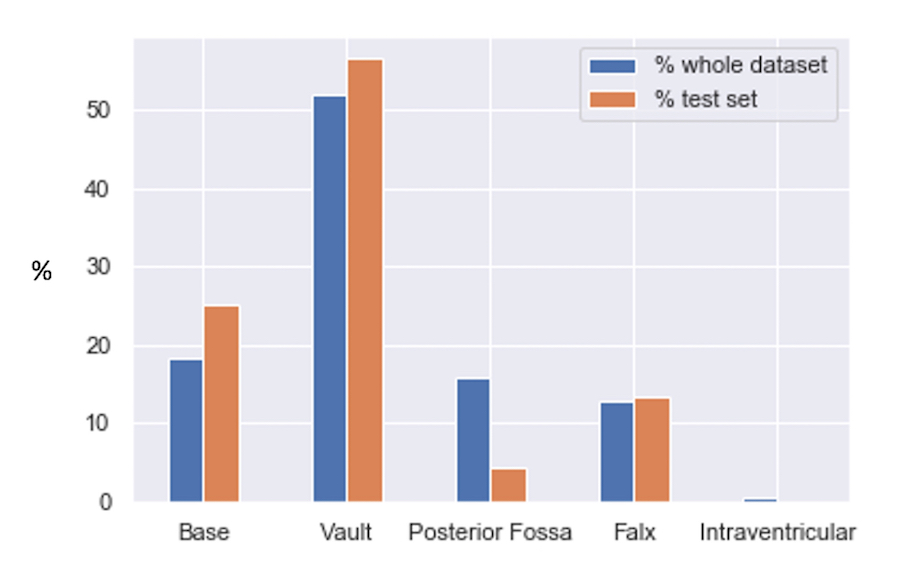

### Supplementary Figure 3

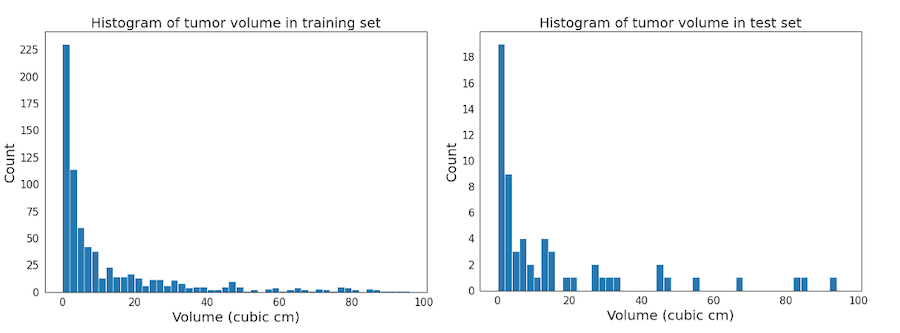

### Supplementary Figure 4

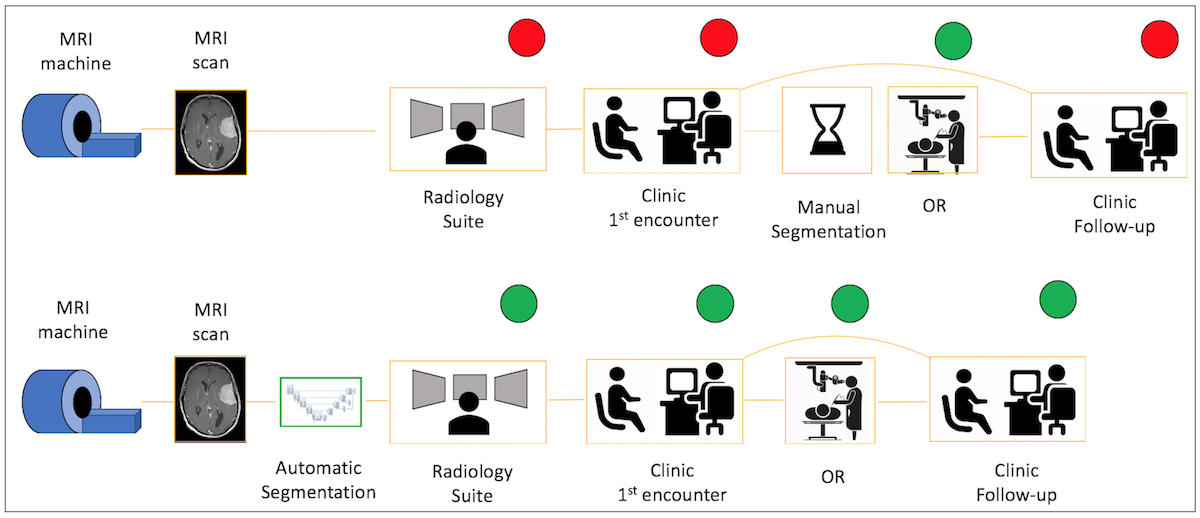
